# Viable virus aerosol propagation by PAP circuit leak and mitigation with a ventilated patient hood – a model for improving health care worker safety in the COVID-19 pandemic

**DOI:** 10.1101/2020.09.04.20187922

**Authors:** Shane A Landry, Jeremy J Barr, Martin I MacDonald, Dinesh Subedi, Darren Mansfield, Garun S Hamilton, Bradley A. Edwards, Simon A Joosten

**Author notes:** **Addresses:** Shane Landry – Monash University BASE facility, 264 Ferntree Gully Road, Notting Hill, 3168 Jeremy Barr - 25 Rainforest Walk, School of Biological Sciences, Clayton Campus, Monash University, Clayton, 3800 Martin I MacDonald - Monash Lung and Sleep, 246 Clayton Rd, Clayton, 3168 Dinesh Subedi - 25 Rainforest Walk, School of Biological Sciences, Clayton Campus, Monash University, Clayton, 3800 Darren Mansfield – Monash Lung and Sleep, 246 Clayton Rd, Clayton, 3168 Garun S Hamilton - Monash Lung and Sleep, 246 Clayton Rd, Clayton, 3168\ Bradley A. Edwards - Monash University BASE facility, 264 Ferntree Gully Road, Notting Hill, 3168 Simon A Joosten – Monash Lung and Sleep, 246 Clayton Rd, Clayton, 3168. **Correspondence:** Author: Shane Landry, PhD, Address: Sleep and Circadian Medicine Laboratory, Ground Floor, Monash University BASE facility, 264 Ferntree Gully Road, Notting Hill, 3168, Victoria, Australia.

## Abstract

**Background:** Nosocomial transmission of SARS-CoV-2 has been a major cause of morbidity and mortality in the COVID-19 pandemic. Emerging evidence suggests patients auto-emit aerosols containing viable respiratory viruses. These aerosols could be further propagated when patients undergo certain treatments including continuous positive airway pressure (PAP) therapy. This study aimed to assess the degree of viable virus propagated from mask leak in a PAP circuit and the mitigation of virus propagation by an air filter combined with a plastic canopy.

**Methods:** Bacteriophage PhiX174 (10^8^copies/mL) was nebulised into a custom PAP circuit within a non-vented clinical room. Mask leak was systematically varied to allow 0, 7, 21, 28 and 42 L/min at the mask interface. Plates containing *Escherichia coli* host assessed the degree of viable virus (via plaque forming unit) settling on surfaces around the room. In order to contain virus spread, the efficacy of a simple, low-cost ventilated headboard, created from a plastic tarpaulin hood and a high efficiency particulate air (HEPA) filter was tested.

**Findings:** Increasing mask leak was associated with virus contamination in a dose response manner (χ^2^ = 58.24, df = 4, p< 0.001). Clinically relevant levels of leak (≥21 L/min) were associated with virus counts equivalent to using PAP with a standard vented mask. The highest frequency of viruses was detected on surfaces 1m from the leak source, however, viable viruses were recorded on all plates (up to 3.86m from source). A plastic hood with HEPA filtration significantly reduced viable viruses on all plates. HEPA exchange rates of 170 and 470m^3^/hr eradicated all evidence of virus contamination.

**Interpretation:** Mask leak from PAP circuits may be a major source of environmental contamination and nosocomial spread of infectious respiratory diseases. Subclinical levels of leak should be treated as an infectious risk. Cheap and low-cost patient hoods with HEPA filtration are an effective countermeasure.

**Funding:** National Health and Medical Research Council of Australia (1139745).

**RESEARCH IN CONTEXT:** *Evidence before this study:* Nosocomial spread of SARS-CoV-2 results in increased infection rates among healthcare workers compared to the general population. Those workers involved in the delivery of non-invasive ventilation are at higher risk based on evidence from previous SARS outbreaks. However, little is known about virus aerosol spread and environmental contamination from respiratory interventions like non-invasive ventilation, which is one of few life-saving treatments for COVID-19 patients. We therefore searched through PubMed with no language restrictions from inception to August 21, 2020 using the search terms ([NIV] or [non-invasive ventilation] or [noninvasive ventilation] or [CPAP] or [continuous positive airways pressure] or [PAP] or [positive airways pressure]) and ([aerosol spread] or [aerosol dispersion] or [aerosol generation]). The search returned 130 publications of which 28 related to the generation or spread of aerosols. Of the 28 related papers, 17 were consensus or opinion papers, 4 were reviews and 7 were original research papers. All previous studies investigating aerosol propagation with respiratory interventions utilised particle sizers or smoke visualisation techniques. These methodological limitations mean that particles are counted or visualised close to the aerosol source and reveal little about wider aerosol spread. Furthermore, they ignore the inherent biological aspects of viral aerosol dispersion in that the aerosol needs to contain viable virus in order to be infectious. It has not been directly established that clinical respiratory interventions are capable of propagating viable virus aerosol and no attempt has been made to systematically quantify the degree of environmental contamination from viable virus aerosol escaping from non-invasive ventilation circuits. There are no current studies informing us as to the effectiveness of air filtration interventions at mitigating environmental contamination with viable virus aerosol escaping from non-invasive ventilation circuits.

*Added value of this study:* Our study quantifies the degree of viable virus aerosol spread from clinically relevant levels of noninvasive ventilator circuit mask leak, and demonstrates a risk mitigation strategy using a hood and air-purifier at completely eliminating viable virus aerosol environmental contamination. We developed a viable virus aerosol model utilising bacteriophage PhiX174 which is similar in size to SARS-CoV-2 and is harmless to humans. Through nebulising a solution of PhiX174 into a custom ventilation circuit with controllable mask leak settings, we were able to demonstrate that increasing circuit leak was associated with environmental virus contamination in a dose response manner (p< 0.001). Even sub-clinically apparent levels of circuit leak (< 7L/min) were associated with detectable virus propagation up to 3.86 metres from the leak source. Deployment of a hood and air-purifier setup as described by the United States Centres for Disease Control and Prevention, completely eliminated environmental virus contamination from viral aerosol dispersion.

*Implications of all the available evidence:* Non-invasive ventilator circuit mask leak can propagate live virus containing aerosol and can lead to extensive environmental contamination up to 3.86 metres from the leak source, even at levels of leak that would be difficult to detect clinically. This raises important safety considerations for open wards delivering non-invasive ventilatory support and could explain the noted increased risk of nosocomial SARS infections in healthcare workers delivering non-invasive ventilation treatment. Point of emission air filtration with simple hood and air-purifier completely eliminates environmental contamination with viable virus and could be readily deployed to protect health care workers in the COVID-19 pandemic.

## INTRODUCTION

The COVID-19 pandemic has placed enormous pressure on public health and hospital systems across the globe. In the context of the ongoing health disaster, healthcare worker (HCW) furlough, morbidity and mortality has further stretched hospital resources. Those workers in forward facing roles caring for COVID-19 patients are at highest risk (1).

Positive Airway Pressure (PAP), applied either as CPAP or Non-invasive Ventilation (NIV) is a frequently employed and life-saving treatment for patients with COVID-19 (2, 3). Given that NIV usage can propagate patient expired air via exhalation ports (4), respiratory circuits are often modified to include a viral filter that cleans exhaled air (5). However, HCWs attending patients with severe COVID-19 who require PAP remain at increased risk of infection (6), even when adequate personal protective equipment is utilised (7). It is not entirely clear why HCWs caring for patients on PAP are at higher risk. Inadequate protective equipment use (8) and NIV mask leak are clear possibilities – although no data exists on the extent to which mask leak is an environmental contamination risk. Importantly, some degree of unintended mask leak is present in all situations where PAP is applied to a mask, and leak is much more likely when a high degree of pressure/ventilatory support is required.

There is mounting evidence that aerosols containing SARS-CoV-2 play an important role in nosocomial spread of COVID-19 infection. Patients with seasonal coronavirus infection generate virus containing aerosol (9) and SARS-CoV-2 aerosols can remain infectious for at least three and up to 72 hours after generation (10). Even when patients are cared for in purpose built biocontainment units with negative-pressure rooms, SARS-CoV-2 can be detected in the air and on surfaces distant from patients (11, 12) as well as parts of the hospital removed from patient care areas (13). Due to the highly contagious nature of SARS-CoV-2 (14) and the high morbidity and mortality associated with COVID-19 (15–17), any environmental contamination poses a risk to HCWs and other patients. In the absence of negative pressure biocontainment units, the Unites States Centres for Disease Control and Prevention (CDC) recommends the use of ventilated hoods with high efficiency particulate air (HEPA) filtration (18), although the effectiveness of these interventions at reducing HCW and environmental contamination has not been established.

Given the current gaps in knowledge we aimed to quantify the amount of viable virus that is propagated from clinically relevant levels of PAP circuit leak. To accomplish this, we used the surrogate virus PhiX174 (*Microviridae* family), which is a tail-less, icosahedral, non-enveloped, bacteriophage with a linear ssDNA genome. Due to its small size (0.025 μm) and intrinsic stability, PhiX174 is commonly used as a viral aerosol model (19, 20). Finally, we determined if a hood with HEPA filter set to different air flow exchange rates can mitigate environmental spread of viable virus aerosol.

## METHODS

A series of experiments were designed to quantify aerosolised viral propagation of a simulated patient with viral respiratory disease (e.g. COVID-19) undergoing PAP with a non-vented mask in a non-negative pressure hospital room. It was assumed based on current data (9) that such a patient would generate aerosols via breathing (or possibly coughing) whilst receiving PAP treatment.

### Bacteriophage PhiX174

Bacteriophage PhiX174 was used as a surrogate virus due to its size (0.025μm), stability to aerosolization (20), and non-pathogenic nature. PhiX174 was propagated using the bacterial host *E. coli C* ATCC13706 grown in Tryptic Soy Broth (TSB). The lysate was purified following the Phage-on-Tap protocol (21) and resuspended in 1X phosphate-buffered saline (PBS; Omnipur, Gibbstown, NJ, USA). Phage titre was determined by soft agar overlay method. To determine viral aerosolization and spread, a series of soft agar plates containing *E. coli C* bacterial host were left uncovered in the room for set periods of time. Settle plates were then sealed, incubated overnight at 37 °C and viral plaques were enumerated the following day (Figure 1A).

**Figure 1.**
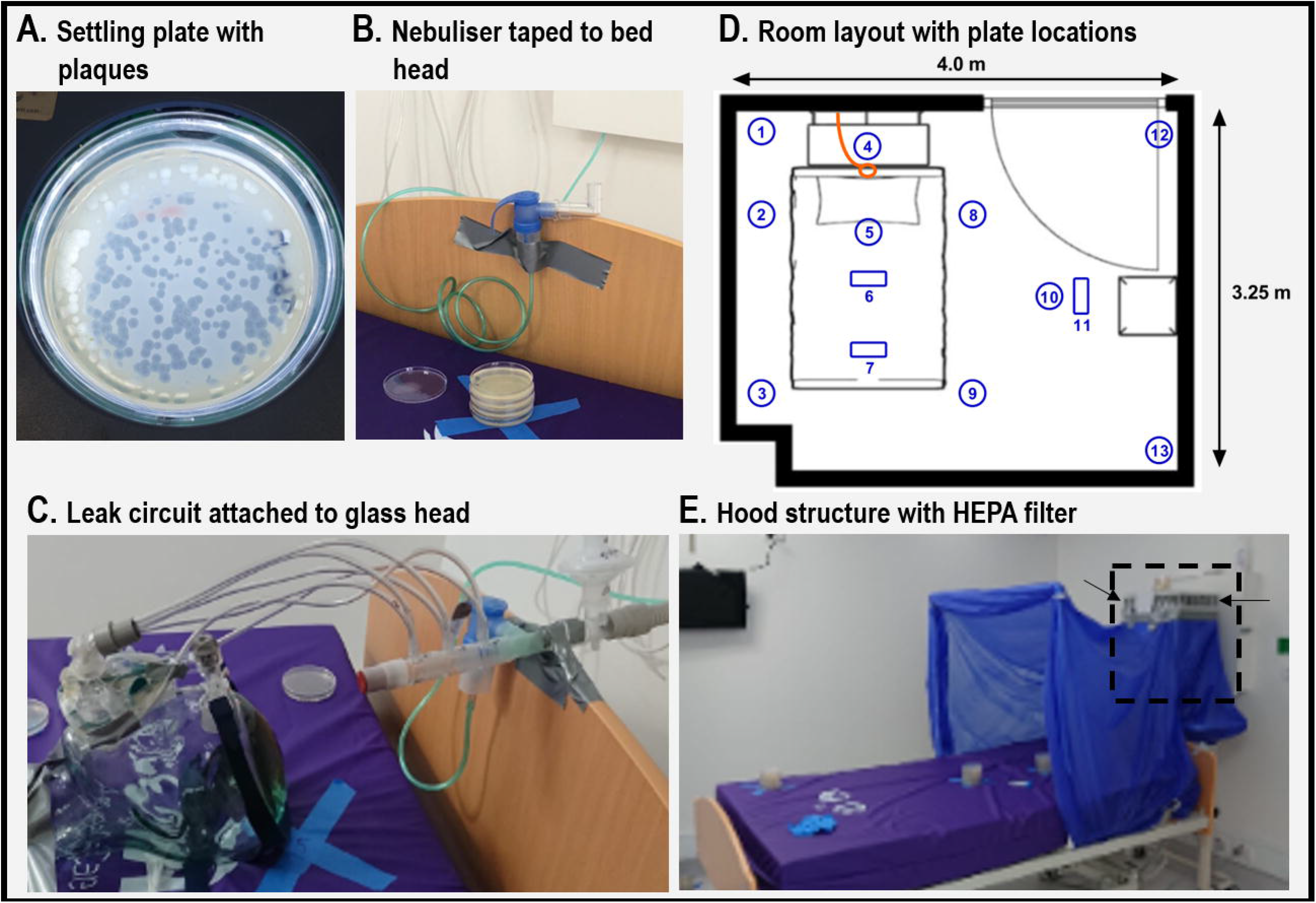
Experimental apparatus. **A.** Photo of a setting plate with visible plaques. Each plaque indicates a single bacteriophage had settled in that precise location. **B**. Nebuliser (filled with 10 mL bacteriophage lysate) taped to bedhead. **C**. Leak circuit (with nebuliser attached), image shows six leak tubes connected to the mask indicating it is configured to produce leak at 42 L/min. **D**. Room layout with locations of 13 numbered plates shown in blue. Circular symbols represent where plates were positioned on floor or bed, rectangular symbols show plates hanging from ceiling oriented perpendicular to the floor. **E**. Hood structure constructed from a plastic tarpaulin sheet and PVC piping is covering the head of the bed. The rear of the hood skirts around HEPA filter (highlighted with black dashed line) and encloses its air intake. Exhaust can be seen just above tape point (see arrow). Details photographs and diagrams are provided in the supplement. See supplementary materials for detailed photographs and diagrams.

A series of detection sensitivity experiments were performed in order to find the optimal phage dose detectable on settling plates. These experiments were informed by known quantities of seasonal coronavirus viral copies emitted by and in the upper airway of ambulant/non-hospitalised patients (9). A nebulised 10 mL solution of 10^8^ PhiX174 virions per mL (for an effective total dose of 10^9^ phages) provided ideal detection sensitivity. Details of the titration experiments can be found in the supplement.

We nebulised a total dose of 10^9^ phages, and Leung et al (9), demonstrated up to 10^5^ seasonal coronavirus copies emitted as aerosol in 30 minutes of breathing, indicating that we should adjust our calculated plaque count by 0.0001 (**10^5^ / 10^9^)** to calculate likely pure aerosol spread of SARS-CoV-2. Further adjustments taking into account i) surface area sampling size and ii) duration of experiment were also carried out (results presented in the supplement).

### Aerosol generation

Aerosols were generated using a nebuliser (Pari-Pep, PARI Respiratory Equipment, VA, USA, Figure 1B). Medical air was delivered at 9 L/min to the nebuliser via tubing connected to a wall mounted flow metre (RTM3 0-15 L/m). The nebuliser was modified by gluing the lid shut to ensure aerosols/airflow could only flow from the ‘mouth-piece’. The Pari-Pep device produces a distribution of aerosol particle size of 3.42 µm ± 0.15 µm (22).

### Simulated circuit leak

A respiratory circuit was created to generate stable and discrete levels of PAP circuit leak (Figure 1C). The circuit comprised a sealed end piece connected to 3 pressure port connectors (Fisher & Paykel, 900HC452) placed in series. These closable pressure ports (6 total) served as ‘leak ports’ in the circuit. Oxygen tubing was connected to each leak port and threaded through the elbow of a PAP mask (Resmed, Quattro) and taped to the edges of the mask. Each tube/port was fixed in place to direct leak towards typical areas of mask leak (corners of the mouth, bridge of the nose, etc). Connected in order from the mask to machine, two T-piece connectors were placed in series, the first connected to the Pari-Pep Nebuliser which served as the aerosol input point. The second T-piece connector (Fisher & Paykel, RT017) was attached to a viral filter (SureGard, RJVKB6, Viral Filtration Efficiency = 99.99%, tested against PhiX174) which served as the filtered expiratory vent. The circuit was attached via CPAP hosing (Fisher & Paykel, 900HC221) to a pressure source (Phillips-Respironics, P_crit_ 3000). CPAP was chosen over bi-level as it delivers a continuous, rather than variable, pressure/leak. Detailed diagrams of the circuit are provided in the supplement (Figures S2 & 3).

CPAP pressure 15.5 cmH_2_O paired with 9 L/min nebuliser air input produced approximately 7 L/min leak increments for each port open such that 0, 7, 21, 28, 42 L/min leak could be generated by opening 0, 1, 3, 4 and 6 leak ports respectively. These leak levels were tested quantitatively and were repeatable (see supplement Table S2) and were chosen because they represent a spread of leak levels likely to be experienced clinically.

### Clinical Room

All experiments were undertaken in a room with dimensions: 4.0 × 3.25 × 2.7 m (surface area 13.0 m^2^, volume 35.1 m^3^) see Figure 1D. All entrances and vents were taped shut to eliminate airflow currents – heating and cooling appliances were switched off. The room was insulated, with continuously recorded temperature (M±SD = 21.4 ± 1.0 ºC) and barometric pressure (758.3 ± 2.5 mmHg) varying minimally during experimental procedures. The room was furnished with a single hospital bed, table, and chair.

Ten settling plates were placed at specific sites around the room to quantify the number of viruses settling on surfaces. Three plates were mounted to hang at head height perpendicular to the floor facing the source. Two settling plates were within 1.0 metre of the bedhead/aerosol source, four plates (3 settling, 1 hanging) were within 1.5 metres, with the remaining plates being between 1.5 metres and 3.86m (exact distances provided in supplementary Table S1).

CPAP hosing and oxygen/air tubing were fed into the room from an external control room (via sealed holes in the wall). The nebuliser and leak circuit were taped to the head of the bed and placed so that the leak outputs were positioned where a patient’s head would normally be (Figure 1C&D).

### HEPA filtration and patient hood structure

An air-purifier with HEPA filter (IQ Air, Healthpro 250) was used to perform multiple air exchanges in order to clear the room of virus between experiments. The air-purifier has 6 flow settings (50, 100, 170, 240, 310, 470m^3^/hr). Pilot testing (see supplementary materials) showed that the room could be adequately cleared by 30 minutes of run time at a flow rate of 470m^3^/hr (approximately 6.7 exchanges).

To test protective measures designed to reduce viral propagation, a hood structure was created modelled on CDC recommendations (18) using hardware store materials (cost ∼$40AUD/$29USD). The structure draped over the top of the bed enclosing the patient’s head and the air intake of the air-purifier (Figure 1E, detailed diagrams provided in the supplement).

### Experimental Protocols

#### Experiment 1

To assess the degree of viable viral aerosol propagation associated with CPAP circuit leak, the bacteriophage lysate was nebulised for 45 minutes directly into the leak circuit, which was pressurised at a constant 15.5 cmH_2_O. Plates were covered and removed at the end of the 45 minute period. This procedure was repeated three times for each of the leak levels (0, 7, 21, 28, 42 L/min). For comparative purposes, an additional condition was run where the viral filter was removed from the expiratory limb of the circuit (equivalent to using a standard vented CPAP mask) and the mask leak was set to 0 L/min. Between each condition the air purifier was run at its highest setting (470m^3^/hr) for 30 minutes. Following this, control plates were placed in the room for 10 minutes to ensure room air was free of virus.

#### Experiment 2

To assess the ability for a protective hood and HEPA filter to reduce aerosolised virus propagation and environmental contamination, the bacteriophage lysate was first nebulised for 30 minutes in the room (unconnected to the CPAP circuit). Plates were covered and replaced at 30, 45, and 60 minutes post-nebulisation by one of the investigators who remained in the sealed room. These conditions were then repeated with the nebuliser placed within the hood, then repeated with the air purifier (within the hood) turned on at 50, 170 and 470m^3^/hr settings.

### Data analysis

In each experiment viable viruses were quantified by counting the number of plaques on settling plates. Plaque counts > 200 were considered too-many-to-count (TMTC) and were rated using an ordinal visual rating scale (+, ++, +++, ++++), with TMTC++++ indicating that complete lysis has occurred on the plate. For graphing and analysis purposes TMTC ratings were given numeric values of 200, 210, 220, and 230. Friedman’s test with post hoc comparisons (Dunn test) was used to compare plaque counts between conditions. A p value > 0.05 was considered statistically significant.

## RESULTS

### Viral aerosol propagation associated with PAP circuit leak

Figure 2 shows the degree of aerosolised virus escaping from the PAP circuit with non-filtered, non-sealed circuit (at 0 L/min leak) as a reference. Increased leak was associated with increase in viral counts across settling plates in a dose-response manner (χ^2^ = 58.24, df = 4, p< 0.001). Post hoc tests showed viral counts were significantly lower in the 0 L/min condition compared to any other leak level. Similarly, viral counts were higher in the 42 L/min leak condition compared to any other level of leak. Mask leak levels ≥21 L/min demonstrated comparable viral counts to when the viral filter was removed from the expiratory vent of the circuit.

**Figure 2.**
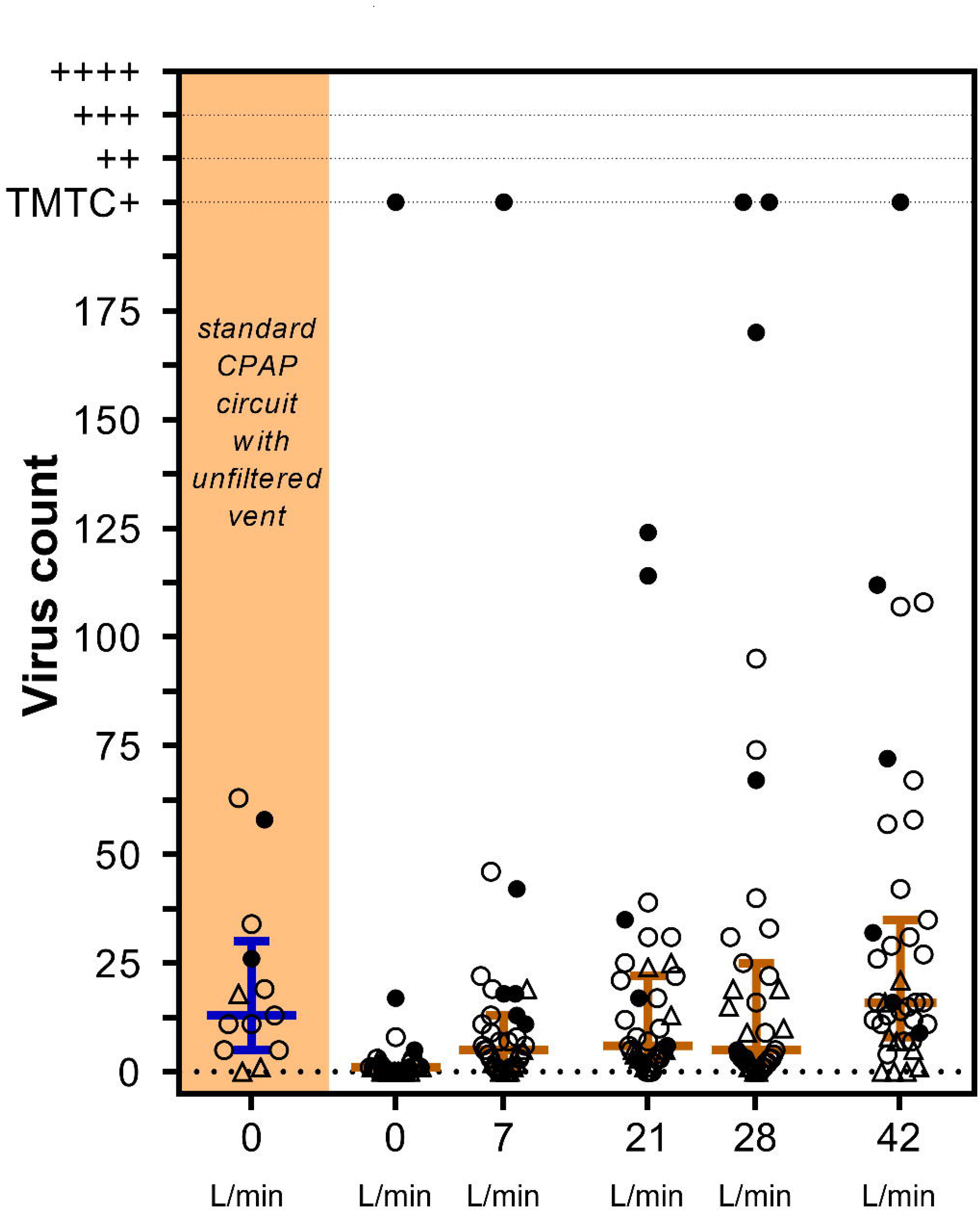
Viral aerosol propagation associated with PAP circuit leak. PhiX174 bacteriophages were nebulised over 45 minutes into in a pressurised (15.5cmH_2_O) PAP circuit designed to leak at either 0, 7, 21, 28 or 42 L/min. As a comparator, the viral filter was removed from expiratory vent and the circuit set to 0 L/min leak (data highlighted in orange). Symbols represent viral counts from settling plates. Solid black circles represent plates 1m from the leak source, triangles represent hanging plates. Orange lines with error bars show median and interquartile range. Virus counts > 200 were considered too-many-to-count (TMTC) and were rated on using an ordinal (+, ++, +++, ++++) visual rating scale.

Plates 4 and 5, which were located less than 1m from the leak point, consistently showed the highest plaque counts and were frequency TMTC. The three hanging plates tended to have lowest plaque counts across all leak levels.

To assess the relationship between viral counts and distance, the viral counts (combined across all leak conditions) were plotted against the distance of each individual plate from the leak source. As shown in Figure 3 there was an inverse monotonic relationship (r_spearmans_ = −0.166, p = 0.02, n = 195) with viral counts decreasing with distance from source. Notably the most distant plate (3.86m) had consistent viral counts at all leak levels ≥7 L/min.

**Figure 3.**
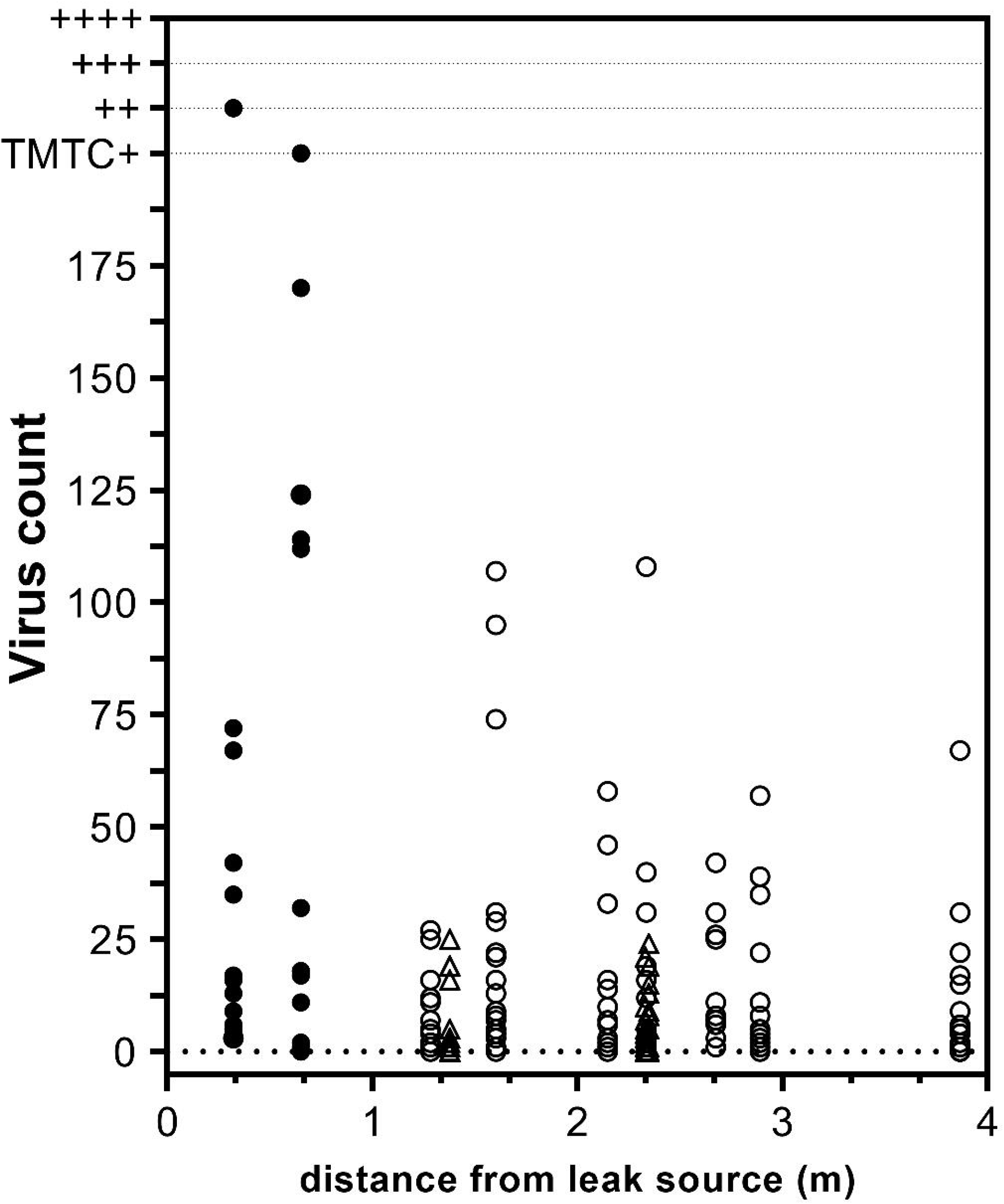
Viral counts versus distance from leak source. Viral counts for each plate, combined across all leak conditions are plotted according to the distance of the plate from the leak source. Specifically, the horizontal and vertical distances of the plate from the leak source were measured and the hypotenuse calculated according to Pythagoras’ theorem. Solid black circles represent plates within 1m from the leak source, triangles represent hanging plates. Virus counts > 200 were considered too-many-to-count (TMTC) and were rated on using an ordinal (+, ++, +++, ++++) visual rating scale

### Efficacy of Hood and HEPA filter

When the bacteriophage solution was nebulised directly into the room without the leak apparatus, high viral counts were found on all settle plates, with significant aerosolised viral load persisting up to 60 minutes after the solution had been completely nebulised (see Figure 4). Comparatively, hanging plates (positions 6, 7 and 11) counts remained relatively low. When viral aerosolization was repeated with the addition of the hood structure, viral counts were significantly attenuated. The further addition of the HEPA filter had minimal additional efficacy at 50m^3^/hr, however at HEPA settings of 170m^3^/hr and 470m^3^/hr all plates registered zero viral counts at all time points.

**Figure 4.**
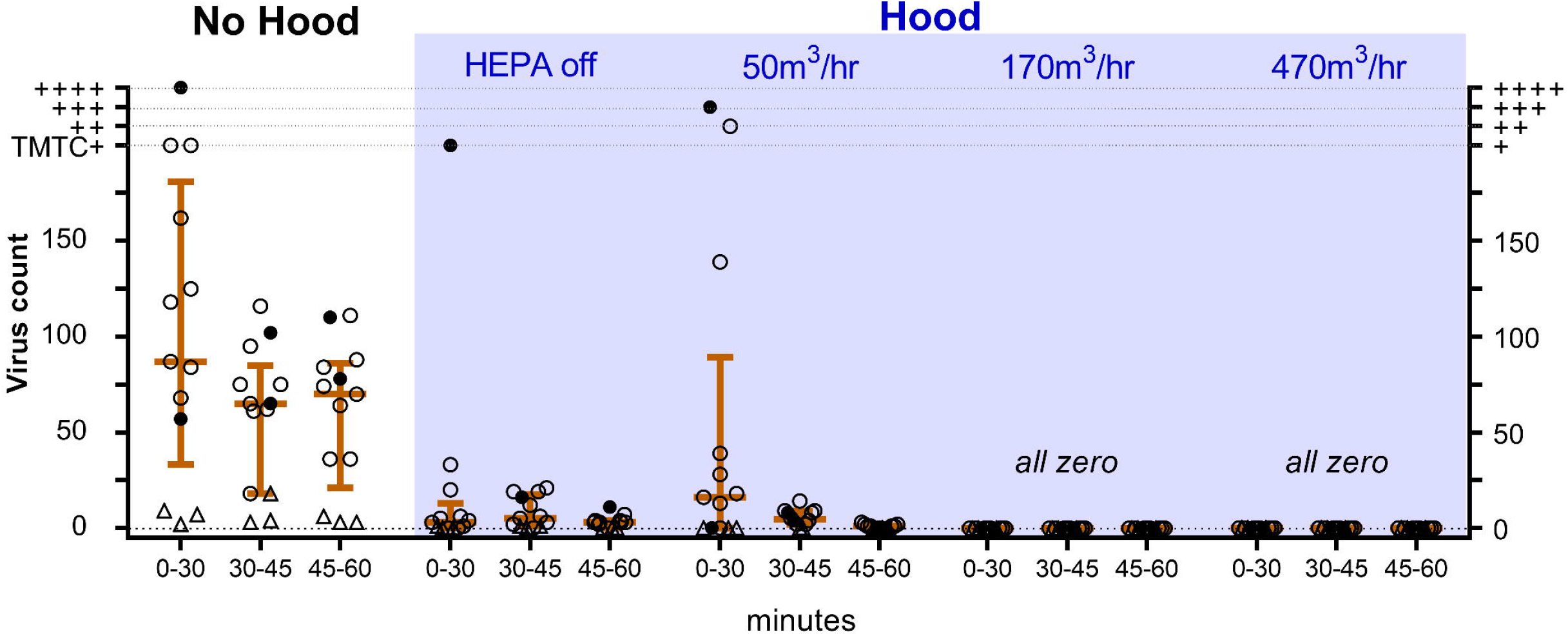
The efficacy of Hood and HEPA filter to reduce viral aerosol spread. PhiX174 bacteriophages were nebulised over 30 mins. Symbols represent plaque counts from settling plates shown across 3-time intervals (0-30, 30-45, 45-60 mins post-nebulisation). A hood structure fitted over the head of the bed substantially attenuated viral counts compared to no hood. No plaques were detected on any plates when HEPA filter was set to 170 or 470m^3^/hr. Solid black circles represent plates within the hood (< 1m from nebuliser), triangle represent hanging plates. Orange lines with error bars show median and interquartile range. Virus counts > 200 were considered too-many-tocount (TMTC) and were rated on using an ordinal visual rating scale.

## DISCUSSION

Using a viable virus model of aerosolised nosocomial transmission, our study quantified the propagation risk associated with unintended PAP system leak, as well as the efficacy of a hood and HEPA filter containment structure to mitigate environmental contamination. Our data shows that aerosols containing viable virus, in similar concentrations to those generated by patients (9), can escape from PAP system leak and can settle onto surfaces at least 3.68m away from the leak source – even at subclinical levels of leak (7 L/min). The degree of leaked virus containing aerosols settling throughout the room is proportional to the amount of leak in a dose response manner. Enclosing the head of the bed in a cheaply constructed hood of plastic sheet and PVC piping substantially attenuated the degree of virus spreading. Moreover, application of a HEPA filter (to the hood) at an exchange rate of 170m^3^/hr or greater, completely eliminated all evidence of virus spreading in the environment, even when 10^9^ viruses were directly nebulised into the air. We believe these findings have immediate and wide-reaching implications for the protection of HCWs on the front line of the COVID-19 pandemic.

Several recent studies have elucidated the risks currently facing HCWs in the COVID-19 pandemic. Firstly, Leung et al (9) demonstrated that ambulatory non-hospitalised patients with seasonal coronavirus are capable of self-generating virus containing aerosol at a rate of up to 10^5^ copies/30mins, in some cases simply by breathing without coughing. We also know that so-called “superspreader” transmission dynamics (where a small proportion of cases are responsible for large numbers of transmissions) are a feature of previous SARS epidemics (23) and the current COVID-19 pandemic (24). The implication is that every COVID-19 patient is a potential silent virus aerosol generator. Secondly, Santarpia et al. (11) demonstrated that even in a dedicated biocontainment facility with negative pressure rooms and hallways, SARS-CoV-2 can be detected on surfaces, including underneath the patient’s bed and at even higher concentrations in air samples from the room and hallway. The presence of such extensive contamination potentially indicates that negative pressure is not absolutely eliminating the route of virus aerosol contamination. Thirdly, van Doremalen et al. (10) demonstrated that SARS-CoV-2 aerosols can remain viable in the environment for at least three and up to 72 hours. These studies help us understand that infected patients generate aerosols which are an important part of extensive environmental contamination even in dedicated specialised environments, and that environmental contamination by aerosols creates potential viable virus risks for HCWs.

Previous studies demonstrated the potential for aerosol propagation from respiratory circuits. Using smoke and lasers to visualise localised aerosol particle spread, Hui et al (4, 25) have shown typically vented CPAP/NIV masks produce (intended) pressure dependent leakage plumes from their exhalation ports in a 1 metre radius. The issue with techniques that visualise particle spread is that whilst they quantify zones of high risk/density environmental contamination, expelled aerosols can travel substantially longer distances than 1 metre where they settle in the environment. Additionally, smoke and particle studies cannot assess the biological aspect of aerosol propagation risk. We have demonstrated that viable virus can be propagated by a respiratory circuit and remain viable in the environment – where it poses a substantial risk for nosocomial transmission. Moreover, virus contained in aerosols were shown to impact on plates at head height, and settle on all surfaces, including the most distant point in the room (3.68 metres from the source). That virus is detectable at distances of greater than 3 metres from the source in our experiments raises important concerns for large open areas such as intensive care units and cohorted wards.

To our knowledge, this is the first study to systematically examine virus aerosol propagation associated with unintended mask leak from a PAP system. Pressurised mask systems are prone to leak that can be difficult to detect at levels < 10 L/min. High pressure requirements, oronasal mask use, coughing, and facial wrinkles/skinfolds are all associated with increased mask leak and are all relevant to COVID-19 patients. Importantly, mask leakage bypasses viral filters placed on the expiratory limb of a PAP circuit. Our study demonstrates that mask leak directly leads to viable virus aerosol propagation in a dose dependent manner, and may suggest that even subclinical levels of leak could be a significant source of risk for HCWs, particularly with prolonged exposure. It is important to note that our experiments quantified the risk associated with short-term use (30-45mins) of PAP, however many patients may require substantially longer periods of use (>24 hours), which multiplies the associated risk.

The CDC predicted that during a pandemic demand for airborne infection isolation rooms would outstrip supply (26). In this context, the CDC has guides for constructing ventilated headboards (18). Several studies have demonstrated efficacy of ventilated hoods at capturing aerosols (27, 28). However, our study is the first to demonstrate the ability of such a structure to eliminate viable virus propagation and environmental contamination. Our study builds on previous data to show even highly contaminated aerosols (containing 10^9^ virus copies) can be eliminated by an apparatus modelled on CDC recommendations. Moreover, we show that this design can be equally effective when constructed from readily available low-cost materials and with modest HEPA air exchanges rates (170m^3^/hr or greater). The fact that we are able to eliminate environmental contamination with modest exchange rates raises important considerations. Santarpia et al (11) showed that even in the absence of AGPs, COVID-19 patients managed in a biocontainment facility with negative pressure at 12 exchanges/hour exhibited extensive environmental contamination including in the air and under the beds, The contrast between a standard negative pressure room and our results at modest air exchanges (170m^3^/hr) is clearly the use of “point of emission” air exchange. We believe the hood structure contains/shepherds aerosol particles and fosters development of a wind tunnel that channels aerosol directly into the air purifier – a feature that is lacking in whole room negative pressure air exchanges.

Our methodological setup has several advantages over previous literature in this field. Bacteriophage PhiX174 has been used in a number of industrial and clinical applications (e.g. testing water and hospital filters). PhiX174 is harmless to humans and is of similar size (∼0.025μm (29)) as SARSCoV-2 (0.060-0.14μm). Using settling plates of *E. coli* enabled us to detect the presence of viable virus with high resolution, in that a single viable copy of the virus causes a visible plaque to be formed on the *E. coli* lawn where the virus has lysed the bacterial host. In this way, our method is an extremely sensitive measure of viable virus propagation and settling in the environment.

This study, however, has several limitations. First, we used a nebuliser which produces a tight range of particle size (3.42µm ± 0.15µm) to produce virus containing aerosols. In contrast, aerosols generated by individuals when they speak or breathe are of similar magnitude (30) but present a much larger range of particle sizes including larger droplet ranges. Larger droplets settle faster and are less likely to travel long distances. Second, because of our detection sensitivity, we aerosolised larger numbers of viruses than has been shown to be emitted as aerosol by infected individuals when breathing (10^9^ vs 10^5^ (9)). However, these levels are well balanced by other factors, such as the degree of surface area sampled within the room and shorter periods of time measured. Further discussion related to the number of viable viruses settling for each given leak is provided in the supplementary materials (Table S3). Importantly, nebulizing 10^9^ phages directly up into the room most likely represents a ‘worse case’ clinical scenario. This showcases the extremely high efficacy of the hood and HEPA filtration structure to mitigate infection risk.

In summary, our results demonstrate that untended mask leak from PAP therapy can be a source of environmental contamination which can be mitigated by the use of a hood and HEPA filter. The hood and portable HEPA filter may represent a relatively low cost and portable adjunct to HCW protection from nosocomial COVID-19 transmission.

## Data Availability

All data referred to in the manuscript is available on request.

## CONTRIBUTORS

SAL, JJB, MIM, SAJ contributed to study concept and design; SAL, JJB, DS, SAJ contributed to acquisition of the data; SAL, JJB, SAJ contributed to data analysis; all authors contributed to initial drafting of the manuscript; all authors contributed to interpretation of the data and critical revision of the report.

## ACKNOWLEDGEMENTS

The authors would like to thank the Monash Cystic Fibrosis Foundation & Dr Christopher Daley for generously donating their air purifier for use in this research. BAE is supported by a Heart Foundation of Australia Future Leader Fellowship (101167). SAJ is supported by a NHMRC early career fellowship (1139745). JJB is supported by NHMRC New Investigator grant (1156588).

